# High rate of major drug-drug interactions of lopinavir-ritonavir for COVID-19 treatment

**DOI:** 10.1101/2020.07.30.20165027

**Authors:** Juan Macías, Ana Pinilla, Francisco A. Lao-Dominguez, Anaïs Corma, Enrique Contreras-Macias, Alejandro González-Serna, Antonio Gutierrez-Pizarraya, Marta Fernández-Fuertes, Ramón Morillo-Verdugo, Marta Trigo, Luis M. Real, Juan A. Pineda

**Author notes:** Corresponding author: Juan Macías. Infectious Diseases and Microbiology Unit, Hospital Universitario Virgen de Valme. Avda Bellavista s/n. 41014-Seville. Spain.

## Abstract

The impact of drug-drug interactions (DDI) between ritonavir-boosted lopinavir (LPV-r) to treat patients with coronavirus disease 2019 (COVID-19) and commonly used drugs in clinical practice is not well-known. Thus, we evaluated the rate and severity of DDI between LPV-r for COVID-19 treatment and concomitant medications. This was a cross-sectional study including all individuals diagnosed of SARS-CoV-2 infection treated with LPV-r and attended at a single center in Southern Spain (March 1^st^ to April 30^th^, 2020). The frequency [95% confidence interval (95% CI)] of potential and major DDI were calculated. Overall, 469 patients were diagnosed of COVID-19, 125 (27%) of them were prescribed LPV-r. LPV-r had potential DDI with concomitant medications in 97 (78%, 95% CI: 69%-85%) patients, and in 33 (26%, 95% CI: 19%-35%) individuals showed major DDI. Twelve (36%) patients with major DDI and 14 (15%) individuals without major DDI died (p=0.010). After adjustment, only the Charlson index was independently associated with death [adjusted OR (95% CI) for Charlson index ≥5: 85 (10-731), p <0.001]. LPV-r was discontinued due to side effects in 31 (25%) patients. Management by the Infectious Diseases Unit was associated with a lower likelihood of major DDI [adjusted odds ratio (95% CI): 0.14 (0.04-0.53), p=0.003). In conclusion, a high frequency of DDI between LPV-r for treating COVID-19 and concomitant medications was found, including major DDI. Patients with major DDI showed worse outcomes, but this association was explained by the older age and comorbidities. Patients managed by the Infectious Diseases Unit had lower risk of major DDI.

## Introduction

Since December 2019, the severe acute respiratory syndrome coronavirus 2 (SARS-CoV-2) infection has become pandemic in a few months. The rapid spread of this novel coronavirus has been further complicated by a wide spectrum of SARS-CoV-2 disease (COVID-19), spanning asymptomatic or mild disease to multiorgan failure ^1–3^. Lacking efficacy-proved treatments, the approach to the management of COVID-19 has been largely empirical, based on indirect data on the antiviral activity of compounds not formally assessed in randomized controlled clinical trials for safety and efficacy ^4,5^.

Ritonavir-boosted lopinavir (LPV-r) was initially used as antiviral for SARS-CoV-2 infection based on indirect data. Thus, LPV-r has in vitro inhibitory activity against severe acute respiratory syndrome coronavirus (SARS-CoV) ^6^. In addition, a non-randomized study suggested a positive clinical and virological effect of LPV-r added to ribavirin among patients with SARS-CoV infection ^7^. LPV-r showed in vitro and in an animal model activity against Middle East respiratory syndrome coronavirus (MERS-CoV) ^8,9^. Viral clearance was reported for some MERS-CoV cases receiving combinations including LPV-r ^10^. Finally, a clinical trial randomizing patients with COVID-19 to standard of care or standard of care plus LPV-r has been reported ^11^. The median time to clinical improvement was shorter in the group treated with LPV-r in the modified intention-to-treat analysis ^11^. However, the trial failed to find differences in the primary objective.

One issue with LPV-r is the potential for drug-drug interactions (DDI) affecting a wide variety of commonly used medications ^12^. Among those, lipid lowering drugs, antiarrhythmics, cardiovascular drugs, analgesics or anticoagulants are frequently used by aged patients. These older patients are more likely to present with more severe COVID-19 requiring admission and respiratory support, and, due to this, more likely candidates to receive drugs with potential antiviral activity against SARS-CoV-2, as LPV-r. Management of LPV-r by non-infectious diseases doctors, not used to the DDI profile of this drug, along with the need for a fast response and the overwhelming burden of COVID-19 in some areas might have led to a high rate of major DDI between LPV-r and concomitant prescriptions. DDI could have an impact on the tolerability of LPV-r and DDI may even be associated with worse outcomes. However, to our knowledge, there is no data on the frequency and impact of DDI between LPV-r to treat patients with COVID-19 and commonly used drugs in clinical practice. Because of these, we evaluated the rate and severity of DDI between LPV-r for COVID-19 treatment and concomitant medications.

## Results

### Characteristics of the study population

Four hundred and sixty-nine patients had a confirmed diagnosis of SARS-CoV-2 infection during the study period. Among them, 125 (27%) individuals were prescribed LPV-r for COVID-19. The median (Q1-Q3) duration of LPV-r treatment was 5 (3-7) days, ranging from 1 to 14 days. Most patients presented comorbidities detected before SARS-CoV-2 infection (Table 1). Thirty-four (27%) patients did not show any previous comorbidity. The median (Q1-Q3) Charlson index was 3 (1-4). Approximately one quarter of the study population showed a Charlson index ≥5 (Table 1). The most frequent clinical presentation of LPV-r-treated patients was pneumonia (Table 1). One hundred and twenty-one (97%) patients were admitted to the hospital, 14 (12%) of them required intensive care, and 26 (21%) died. The characteristics of the study population are summarized in table 1.

**Table 1.** Characteristics of the patients (n=125)

| Characteristic | Value |
| --- | --- |
| Median (Q1-Q3) age, years | 63 (53-76) |
| Male sex, n (%) | 60 (48) |
| Health care professional, n (%) | 14 (11) |
| Nursing home residency, n (%) | 4 (3.2) |
| Comorbidities*, n (%) |  |
| Arterial hypertension | 25 (20) |
| Type 2 diabetes mellitus | 11 (8.8) |
| Cardiovascular disease | 18 (14) |
| Chronic renal disease | 4 (3.2) |
| Chronic lung disease | 6 (4.8) |
| Cirrhosis | 2 (1.6) |
| Dementia | 14 (11) |
| Cancer | 7 (5.6) |
| Drug-related immunosuppression | 3 (2.4) |
| HIV infection | 1 (0.8) |
| Charlson index, n (%) |  |
| 0 | 15 (12) |
| 1-2 | 47 (38) |
| 3-4 | 37 (30) |
| ≥5 | 26 (21) |
| COVID-19 clinical presentation, n (%) |  |
| Extrapulmonary infection | 15 (12) |
| Pneumonia | 96 (77) |
| Severe pneumonia | 12 (9.6) |
| Acute severe respiratory distress syndrome | 2 (1.6) |
| SARS-CoV-2 diagnosis, n (%) |  |
| PCR | 122 (98) |
| Serology | 3 (2.4) |

\*Main comorbidity per patient: For each patient a single comorbidity was selected, i.e. the condition considered most severe.

### Frequency and severity drug-drug interactions

Overall, LPV-r had DDI with concomitant medications in 103 (82%, 95% CI: 75%-89%) subjects. In 97 (78%, 95% CI: 69%-85%) patients, potential DDI were disclosed, and in 33 (26%, 95% CI: 19%-35%) individuals major DDI were found. Concomitant medications with potential DDI classified by therapeutic group are summarized in figure 1a. Drugs with major DDI with LPV-r are shown in Figure 1b. Twenty-two (23%) of 97 individuals undergoing potential DDI between LPV-r and their concomitant medication died compared with 4 (14%) of 28 patients without potential DDI (p=0.335). Twelve (36%) of 33 patients with major DDI and 14 (15%) of 92 individuals without major DDI died (p=0.010). After adjustment, only the Charlson index was independently associated with death (Table 2).

**Table 2.**
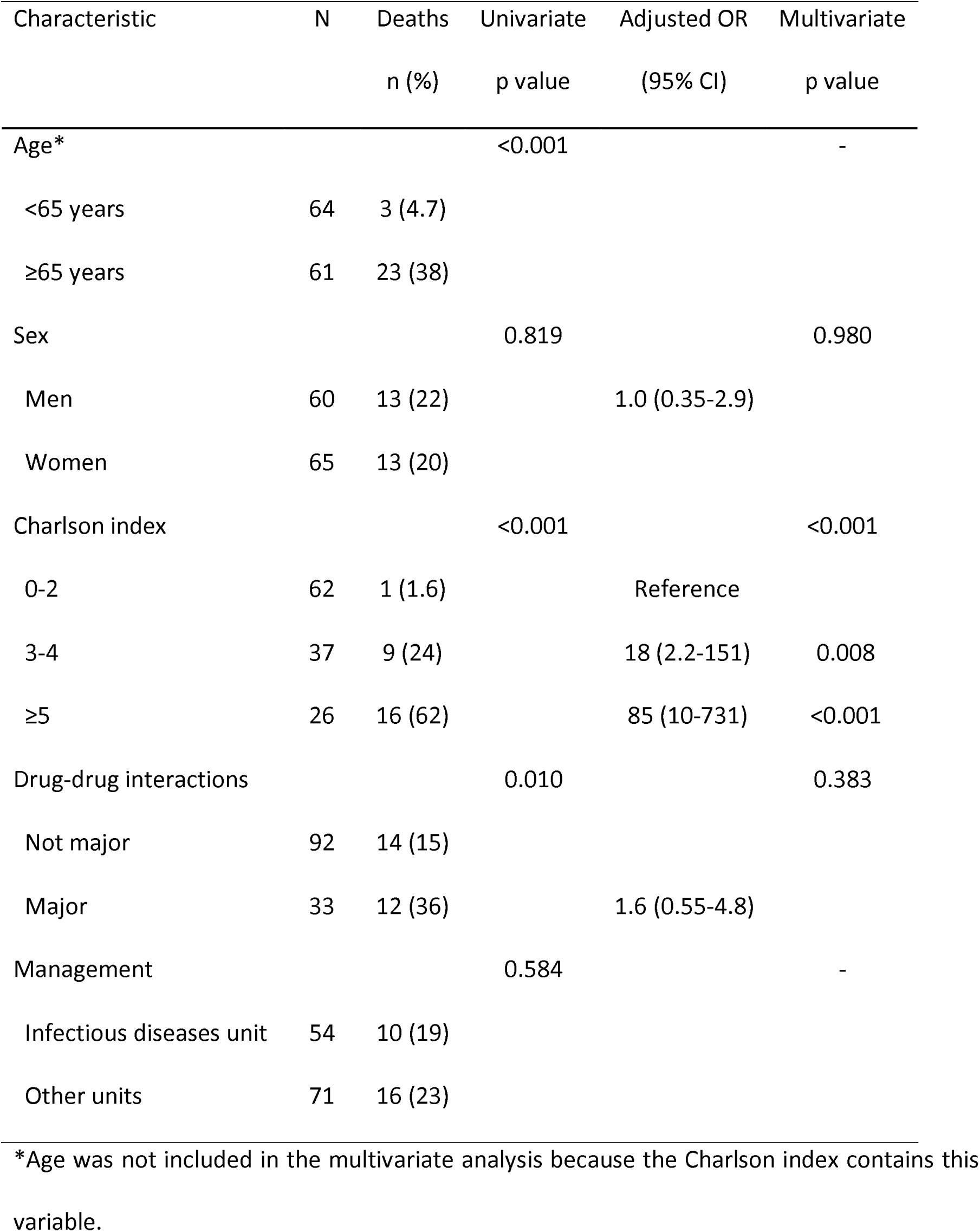
Factors associated with death among patients with COVID-19 receiving lopinavir-ritonavir (n=125).

Among patients managed in the Infectious Diseases Unit, 41 (76%) of 54 individuals showed potential DDI compared with 56 (79%) of 71 patients treated outside the Infectious Diseases Unit (p=0.695). Major DDI were observed in 9 (17%) patients vs. 24 (34%) patients managed by the Infectious Diseases Unit vs. units other than the Infectious Diseases Unit, respectively (p=0.031). The median Charlson index was 2 (1-4) compared with 3 (2-4) for patients managed by the Infectious Diseases Unit and by other units, respectively (p=0.024). The proportion of patients with major DDI treated within the Infectious Diseases Unit by the Charlson index was: Charlson index 0-2, 2 (6.1%); Charlson index 3-4, 1 (8.3%); Charlson index ≥5, 6 (67%) (p<0.001). The proportion of patients with major DDI managed by other units by the Charlson index was: Charlson index 0-2, 7 (24%); Charlson index 3-4, 11 (44%); Charlson index ≥5, 6 (35%) (p=0.303). After adjustment by sex and the Charlson index, management by the Infectious Diseases Unit was independently associated with a lower likelihood of major DDI [adjusted odds ratio (95% CI): 0.14 (0.04-0.53), p=0.003).

LPV-r was discontinued due to side effects in 31 (25%) patients. One patient with pre-existing chronic end-stage renal failure suffered grade 3 diarrhea. LPV-r was discontinued, but renal failure worsened, and the patient ultimately died. Another patient with cardiovascular disease was treated with amiodarone for cardiac arrhythmia. He presented an orthostatic syncope, and LPV-r was discontinued, but he died of sudden death afterwards.

## Discussion

In the present study, we found a high overall frequency of DDI between LPV-r for treating COVID-19 and concomitant medications. Moreover, nearly one third of the patients were treated with commonly used medications with major DDI with LPV-r. Patients in whom major DDI were observed showed a worse clinical outcome, but this association was explained by the older age and comorbidities of these patients. Finally, patients managed by the Infectious Diseases Unit had lower risk of major DDI.

Coadministration of LPV-r with drugs that are major substrates of and highly dependent on CYP3A for clearance is associated with increased plasma concentrations of such drugs or their metabolites. In our study, many commonly used medications with that metabolic interaction with LPV-r were prescribed to patients with COVID-19. Antiarrhythmics, symvastatin or budesonide were found to be administered to the study patients. Elevated plasma concentrations of some of those medications are associated with life-threatening reactions. In this regard, we found a significantly higher frequency of deaths among patients who were treated with medications with major DDI with LPV-r. However, after adjustment in a multivariate model, drugs with major DDI with LPV-r were not independently associated with the risk of death. Instead, the main predictor of death was the Charlson index. Therefore, the patient profile, i.e. older individuals with concomitant diseases needing drugs posing contraindications with LPV-r, accounted for the association between deaths and major interactions with LPV-r.

Patients with COVID-19 treated with LPV-r in the present study were severely ill. The majority needed admission and had high Charlson index scores. More importantly, the overall death rate was 21%, i.e. greater than the mortality reported in hospitalized patients in China ^1,2^. The older age of patients and their elevated Charlson index could explain the higher death rate in our study.

Discontinuations of LPV-r due to adverse events among HIV-infected patients were lower than 10% after two years of antiretroviral treatment ^14^. In a randomized clinical trial of LPV-r for COVID-19 in China, patients stopped early a course of 14 days of LPV-r even more frequently ^11^. Nearly 14% of LPV-r recipients were unable to complete treatment due to adverse events in that trial. In our study, one fourth of the patients discontinued LPV-r because of severe adverse events. The aged and fragile population receiving LPV-r in our study could be more prone to develop LPV-r-related side effects. In fact, our study population was older and accumulated more coexisting conditions than the patients recruited in the Chinese trial of LPV-r for COVID-19 ^11^. The elevated frequency of LPV-r discontinuations in fragile patients with severe COVID-19 questions the clinical applicability of LPV-r as antiviral in such patients.

We found that major DDI between comedications and LPV-r were more frequent among patients managed by non-Infectious Diseases Units compared with the Infectious Diseases Unit. After adjustment by the Charlson index, an independent association between major DDI with LPV-r and the treating unit was confirmed. A few factors could explain this finding. First, the overwhelming COVID-19 crisis in our center led to admitting patients with COVID-19 in wards other than the Infectious Diseases ward. In this situation, treatment stewardship by Infectious Diseases doctors was not feasible. Second, management by Infectious Diseases specialists has demonstrated to improve the outcome of patients with other infections ^15,16^. However, even Infectious Diseases physicians, more aware of the DDI of ritonavir-boosted HIV protease inhibitors than other specialists, exposed patients to major DDI with LPV-r. The complexity of patients, staff shortage due to quarantines of doctors falling ill or exposed to unprotected close contacts, and the many difficulties imposed by a health crisis might partly explain those prescription mistakes.

The present study may have some limitations. First, due to LPV-r supply shortage, this antiviral drug was restricted mostly to more severe patients, those requiring hospitalization. This population represents an extreme of the pandemic, with more complications needing medical intervention and worse outcomes. LPV-r given to younger and less fragile patients might have been more tolerable, with less DDI and discontinuations. Second, post-discharge deaths at home or nursing facilities were not evaluated, only deaths during admissions or at the emergency room were evaluable for the present study. Death rates could have been even higher than those reported herein with a follow-up after discharge, as suggested by another study ^17^. Third, the main source of data was the electronic clinical records and the Pharmacy electronic prescriptions. This might have precluded detailed medical record review and it could have led to data loss. However, currently in the Andalusian Health System, health and medical records of every patient are collected in a common electronic database by all caring physicians and nurses. Medications are also prescribed using electronic prescriptions both for outpatients and inpatients. In this regard, individual patient electronic medical records were manually evaluated. These are strengths of the present study.

In conclusion, LPV-r for hospitalized patients with COVID-19 frequently interacts with concomitant medications. Major DDI contraindicating the coadministration of LPV-r and common medications were alarmingly overlooked in the setting of the COVID-19 health care crisis. Safer antiviral treatments for COVID-19, with a better interaction profile, are clearly warranted.

## Patients and methods

This was a cross-sectional study. All individuals diagnosed of SARS-CoV-2 infection from March 1^st^ to April 30^th^, 2020 attended at a single center in Seville, Southern Spain, were evaluated to identify those treated with LPV-r. All patients with COVID-19, confirmed by SARS-CoV-2 PCR or serology, who were prescribed LPV-r as a part of their treatment for SARS-CoV-2 infection were included. The electronic clinical records and Pharmacy electronic prescriptions of all patients were examined to select those who received at least one dose of LPV-r. Concomitant medications were collected from the electronic prescriptions. The electronic clinical records were used to identify comorbid conditions to calculate the Charlson index for each patient ^13^. The vital status of patients was established querying electronic clinical records and it was last updated in May 22^nd^, 2020.

DDI were evaluated using the COVID-19 Drug Interactions site (https://www.covid19-druginteractions.org/) by the Liverpool Drug Interaction Group. The frequency and grade of DDI was assessed. DDI were graded, according to the Liverpool Drug Interaction Group, as potential and major. Potential DDI does not preclude co-administration, since they are usually manageable, but they indicate the need to balance risks and benefits for an individual patient. Major DDI correspond to the “do not co-administer” classification of the Liverpool Drug Interaction Group. The death rate by the severity of DDI was assessed. In addition, side effects leading to discontinuation of LPV-r were examined.

### Statistical analysis

Continuous variables were compared by the Student-t test, or the Mann-Whitney U if applicable. Categorical variables were compared using the Chi-square test, or the Fisher test where necessary. 95% confidence intervals (95% CI) were calculated for the overall, potential and major frequencies of DDI. Factors associated with death were entered in a logistic regression model, adjusted by age and sex. The STATA 16.0 and IBM SPSS 25 packages were used for the analysis.

### Ethics issues

The study was designed and performed according to the Helsinki declaration and approved by the Ethics Committee of the Valme University Hospital (Seville, Spain). Informed consent was waived by the Ethics Committee of the Valme University Hospital due to the observational retrospective design of the study.

## Data Availability

All data is reported in the paper

## Data availability

All data generated or analyzed during this study are included in this article.

## Acknowledgements

The authors wish to thank the collaboration of the nurse M<u>^a^</u>Victoria Martínez.

This study has not been funded. J.M. is the recipient of a grant from the Servicio Andaluz de Salud de la Junta de Andalucía (grant number B-0037). J.A.P. is recipient of an intensification grant from the Instituto de Salud Carlos III (grant number Programa-I3SNS).

## Author contributions statement

Study concept and design: JM, JAP, LMR, RMV

Acquisition of data: AP, FLD, AC, ECM, AGS, AGP, MFF, MT

Analysis and interpretation of data: JM, JAP, LMR

Drafting of the manuscript: JM, JAP, LMR, RMV

Critical revision of the manuscript: All authors.

Final approval: All authors

---

Figure 1. **A.** Proportion of patients with potential drug-drug interactions by concomitant medication group (N=125). GI drugs: gastrointestinal drugs included ondansetron, metoclopramide and loperamide. **B.** Proportion of patients with major drug-drug interactions by concomitant medication (N=125). Other drugs: ivabradine, ticagrelor, domperidone, quetiapine.

